# Magnesium Treatment on Methylation Changes of Transmembrane Serine Protease 2 (TMPRSS2)

**DOI:** 10.1101/2021.03.11.21253287

**Authors:** Lei Fan, Xiangzhu Zhu, Yinan Zheng, Wei Zhang, Douglas L Seidner, Reid Ness, Harvey J Murff, Chang Yu, Xiang Huang, Martha J Shrubsole, Lifang Hou, Qi Dai

## Abstract

**Background:** The viral entry of SARS-CoV-2 requires host-expressed TMPRSS2 to facilitate the viral spike (S) protein priming.

**Objectives:** To test the hypothesis that Mg treatment leads to DNA methylation changes in *TMPRSS2*.

**Methods:** This study is nested within the Personalized Prevention of Colorectal Cancer Trial (PPCCT), a double-blind 2×2 factorial randomized controlled trial, which enrolled 250 participants from Vanderbilt University Medical Center. Target doses for both Mg and placebo arms were personalized.

**Results:** We found that 12-week of personalized Mg treatment significantly increased 5-mC methylation at cg16371860 (TSS1500, promoter) by 7.2% compared to placebo arm (decreased by 0.1%) in those aged < 65 years old. The difference remained statistically significant after adjusting for age, sex and baseline methylation as well as FDR correction (FDR-adjusted *P* =0.014). Additionally, Mg treatment significantly reduced 5-hmC level at cg26337277 (close proximity to TSS200 and 5’UTR, promoter) by 2.3% compared to increases by 7.1% in the placebo arm after adjusting for covariates in those aged < 65 years old (*P*=0.003). The effect remained significant at FDR of 0.10 (adjusted *P* value=0.088).

**Conclusion:** Among individuals aged younger than 65 years with the Ca:Mg intake ratios equal to or over 2.6, reducing Ca:Mg ratios to around 2.3 increased 5-mC modifications (i.e. cg16371860) and reduced 5-hmC modifications (i.e. cg26337277) in the *TMPRSS2* gene. These findings, if confirmed, provide another mechanism for the role of Mg intervention for the prevention of COVID-19 and treatment of early and mild disease by modifying the phenotype of the *TMPRSS2* genotype.

## INTRODUCTION

As of December 4^th^ 2020, in the US alone, coronavirus disease 2019 (COVID-19) caused by severe acute respiratory syndrome coronavirus 2 (SARS-CoV-2), led to 14.2 million confirmed cases and 276,000 deaths. Up to 20% of symptomatic individuals will progress to severe or critical illness ranging from hospitalization to death while some with mild symptoms may also experience a variably prolonged period of recovery with long-term complications [1]. Current antiviral pharmaceutical therapeutics, such as remdesivir, targeting hospitalized patients with COVID-19 have not achieved statistically significant benefits on mortality in randomized trials [2–5]. While awaiting global vaccination for SARS-CoV-2 to end the COVID-19 pandemic, and confirmation that vaccination provides complete protection among adults and for multiple SARS-CoV-2 variants [6–8], the National Institute of Health (NIH) highlights an urgent need for interventions which can be administered early during the course of infection to prevent disease progression to severe COVID-19, speed recovery, and prevent long-term complications [1].

In a precision-based randomized trial [9], we reported that an improved magnesium (Mg) status led to an optimal level of vitamin D. A recent trial further confirmed this finding that Mg treatment improved vitamin D status [10]. We also previously found high circulating levels of vitamin D were prospectively related to reduced risk of cardiovascular (CVD) mortality only when Mg intake was adequate [11]. Based on our findings, a recent study conducted by Tan et al. found a combinatorial treatment of Mg and vitamin D_3_ within the Recommended Dietary Allowance (RDA) plus vitamin B12 reduced the risk of severe illness from COVID-19 by 80% [12]. Although cross-sectional studies investigating the correlation between vitamin D levels and COVID-19 severity and mortality [13] were inconclusive and a large-scale prospective cohort study in the UKBiobank found that serum 25(OH)D (25-hydroxyvitamin D) was not related to risk of severe COVID-19 after adjusting for confounding factors[14], a recent open-label trial using calcifediol (i.e. 25(OH)D) treatment starting at 64 times the RDA levels[15] substantially reduced the need for intensive care unit (ICU) care among hospitalized COVID-19 patients [16].

In addition to optimizing 25(OH)D levels, Mg also activates the conversion from 25(OH)D to 1,25(OH)_2_D, the active form of vitamin D [17]. Patients with “Mg-dependent vitamin-D-resistant rickets” [18] caused by Mg deficiency were resistant to vitamin D treatment alone with no change in blood measures of 1,25(OH)_2_D at doses up to 600,000 IU [17]. However, they dramatically responded to Mg treatment, particularly treatment of Mg plus vitamin D [17,19]. Collectively these varied findings support the hypothesis that the synergistic interaction between Mg and vitamin D may substantially reduce the doses required for vitamin D to reduce the severity of COVID-19. Since 79% of US adults do not meet the RDA of Mg [20], thus, using the combinatorial prophylaxis strategy of vitamin D plus Mg could be a crucial strategy for treating COVID-19.

The viral entry of SARS-CoV-2 depends on binding of the viral spike (S) proteins to cellular receptors (i.e. angiotensin-converting enzyme (ACE2)) which requires host-expressed transmembrane serine protease 2 (TMPRSS2) to facilitate S protein priming [21]. *In vitro* studies showed that TMPRSS2 inhibitor partially blocked SARS-CoV-2 from entering into lung epithelial cells [21]. Animal studies found that *TMPRSS2* knockout mice infected with H1N1 influenza had minimal infection and largely attenuated disease severity[22].

Supporting the findings in mice, a recent cohort found that *TMPRSS2* expression levels and genetic variants played an essential role in modulating COVID-19 severity [23]. The effect is also likely through regulating coagulation cascade and arterial thrombosis [24–26], one key factor for COVID-19 severity and mortality [27,28]. *TMPRSS2* expression is influenced by both genetic variants and epigenetic changes such as DNA methylation. 5-methylcytosine (5-mC), namely the methylation of the fifth carbon of cytosines, was shown to be associated with transcription repression [29,30] whereas 5-hydroxymethylcytosine (5-hmC) is specifically enriched in tissue-specific enhancers [31] and critical in maintaining active gene expression [32,33]. DNA methylation changes are inducible by environmental exposures including nutrients[34,35]. In addition to optimizing vitamin D synthesis and metabolism, Mg also affects the metabolism of α-ketoglutarate [36,37], one key factor for the ten-eleven translocation (TET) enzymes [38], which catalyze the oxidation of 5-mC to 5-hmC in an active demethylation pathway [39]. We, therefore, aim to test the hypothesis that Mg treatment leads to DNA methylation changes (i.e. 5-mC and 5-hmC) in *TMPRSS2* in the Personalized Prevention of Colorectal Cancer Trial (PPCCT), a precision-based magnesium supplementation trial.

## MATERIALS and METHODS

### Participants, randomization and blinding

This is an ancillary study nested in the parent study, the “Personalized Prevention of Colorectal Cancer Trial” (**PPCCT**; NCT01105169 at ClinicalTrials.gov). The PPCCT is a double-blind 2×2 factorial randomized controlled trial conducted at Vanderbilt University Medical Center, Nashville, TN. The detailed study design has been reported previously [9,40]. In brief, participants aged 40 to 85 years old were recruited including: 1) 236 individuals with adenomas or hyperplastic polyps diagnosed from 1998 to 2014 or 2) 14 polyp-free individuals with a high risk of colorectal cancer. Dietary and total intakes of Ca, Mg, and the Ca: Mg ratios were derived from six 24-hr dietary recalls over the course of the trial including intakes of Ca and Mg supplements. Eligible participants were those who had a Ca intake ≥ 700 mg/day and < 2000 mg/day and in whom the Ca:Mg intake ratio was ≥ 2.6 based on the average of the first two baseline 24-hr dietary recalls. Eligible participants were randomized to Mg glycinate or placebo (microcrystalline cellulose) capsules. The Mg treatment used a personalized dose of Mg supplementation that would reduce the Ca:Mg intake ratio to around 2.3, suggested by previous studies [9,41–43]. Participants, study investigators and staff were blinded to the assigned interventions. Blood samples were collected and processed at each clinic visit. Anthropometric measurements (weight, height, waist and hip circumference) were measured at least twice at each clinic visit.

265 participants were randomized and allocated to the Mg treatment arm or the placebo arm. 15 participants withdrew consent before taking Mg treatment or placebo. Of these, 250 randomized participants started treatment, and 239 completed the trial with 11 participants finishing part of the study before withdrawing [9]. Six of the withdrawals were due to self-reported adverse events (four withdrawals in the treatment arm and two in the placebo arm). One of them had donated blood at baseline and at the end of the trial. Therefore, in the current study, 240 participants were included who had blood collected at baseline and at the end of the trial.

### Measurement of 5-mC and 5-hmC at single resolution for the *TMPRSS2* gene

All 240 participants who were enrolled in the PPCCT and had blood DNA samples available at the baseline and the end of the trial were included in the current study to examine the effect of personalized Mg treatment on methylation modifications in the *TMPRSS2* gene. In order to minimize between-batch variations, samples were randomly organized in treatment-placebo (i.e. one treatment arm with one placebo arm) sets (4 samples in each set: 2 from pre-, and 2 from post-treatment). Lab staff were blinded to sample status (i.e. treatment vs. placebo and pre-treatment vs. post-treatment status).

Genomic DNA was extracted from buffy coat fractions collected using a QIAamp DNA mini-kit (Qiagen Inc, Germantown, MD) according to the manufacturer’s protocol [44]. DNA quality was examined using standard molecular biology protocols. We used the TET-assisted bisulfite (TAB)-Array, which combines TET-assisted bisulfite conversion with the Infinium Methylation EPIC array (EPIC array) that interrogates ∼850,000 CpG or non-CpG methylation sites to differentiate 5-hmC and 5-mC signals at base resolution [45]. Our detailed approach was reported previously [45–47] Using this technique, we were able to differentiate 5-mC from 5-hmC.

The β-values for 5-mC and 5-hmC were estimated using the Maximum Likelihood Estimate from the paired bisulfite conversion and TAB-treated samples ^39^. The following quality control steps were taken: 1) We excluded low-quality probes where the number of beads < 3 or the detection P-value > 0.05 [48]; 2) Exclusion of CpGs with a detection rate <95% and samples with the percentage of low-quality methylation measurements >5% or extremely low signal of BS probes [48]; 3) Exclusion of extreme outliers, as defined by the Tukey’s method [49], based on average total signal value across CpG probes; 4) Remaining samples were preprocessed using the R package ENmix to improve accuracy and reproducibility [48]; 5) Dye bias was corrected using regression on a logarithm of internal control probes [50]; 6) Quantile-normalization of signal for Infinium I or II probes; and 7) Lastly, extreme β value (i.e., the proportion of methylated signal in total signals from 0 to 1) outliers across samples, defined by Tukey’s method, were set as missing. In total, the TAB-Array data for 224 participants out of 240 passed the seven quality control steps. In the current study, we kept all 32 CpG sites related to the *TMPRSS2* gene selected by the EPIC array for 224 participants whose methylation data passed the quality control steps.

### Statistical analyses

Continuous demographic variables (mean ± standard deviation) and categorical demographic variables (percent) were compared between treatment and placebo arms. The Wilcoxon rank sum test was conducted to evaluate continuous variables, while Pearson chi-squared tests were conducted to compare categorical variables. Linear regression models were fitted to examine the effect of Mg treatment on overall changes of cytosine modifications (5-mC and 5-hmC) in the *TMPRSS2* gene in three models, respectively. Model 1: crude value; model 2: sex and age; model 3: sex, age and baseline methylation. We also conducted stratified analyses by age because elderly adults are at increased risk for severe illness and mortality from COVID-19 [51]. All *P* values are two sided and statistical significance was determined using an alpha level of 0.05. The data analyses used R software (version 3.5.1).

## RESULTS

The baseline demographic characteristics of 240 participants are presented in **Table 1**. The treatment arm was not significantly different from the placebo arm for baseline demographic variables, including age, sex, smoking status, alcohol drinking status, physical activity status, educational achievement, race, daily intake of total energy, total Ca, intake ratio of Ca to Mg and factors related to cardiovascular events, including body mass index (BMI), blood pressure and eGFR (**Table 1**).

**Table 1.** Descriptive characteristics of treatment vs. placebo at baseline

|  | Placebo<br>(N=120) | Treatment<br>(N=120) | *P* value |
| --- | --- | --- | --- |
| Age, year | 61.3±8.2 | 60.3±7.7 | 0.48<sup>1</sup> |
| Sex - male (%) | 51.7 | 54.2 | 0.70<sup>2</sup> |
| Body mass index (BMI), kg/m<sup>2</sup> | 30.6±6.6 | 29.9±6.1 | 0.48<sup>1</sup> |
| Systolic blood pressure (mmHg) | 128.3±14.2 | 126.3±15.4 | 0.31 |
| Diastolic blood pressure (mmHg) | 74.7±9.2 | 75.3±8.2 | 0.65 |
| eGFR, ml/min/1.73m<sup>2</sup> | 78.9±14.9 | 81.7±13.8 | 0.13<sup>1</sup> |
| Smoking status (%) |  |  | 0.27<sup>2</sup> |
| Never | 49.6 | 60.0 |  |
| Ever | 42.0 | 33.3 |  |
| Current | 8.4 | 6.7 |  |
| Drinking status (%) |  |  | 0.24<sup>2</sup> |
| Never | 32.5 | 42.5 |  |
| Ever | 20.8 | 20.0 |  |
| Current | 46.7 | 37.5 |  |
| Physically active in ≥ 2 days per week (%) | 77.5 | 85.0 | 0.14<sup>2</sup> |
| Education under college (%) | 9.99 | 10.0 | 0.41<sup>2</sup> |
| Race (%) |  |  | 0.56<sup>2</sup> |
| White | 99.2 | 98.3 |  |
| Daily nutrients intake |  |  |  |
| Total energy (kcal) | 2108.2± 604.5 | 2084.3±547.0 | 0.62<sup>1</sup> |
| Total Ca (mg) | 1251.0±358.6 | 1299.8± 332.0 | 0.20<sup>1</sup> |
| Total Mg (mg) | 337.7± 98.7 | 363.8±96.7 | 0.03<sup>1</sup> |
| Ca:Mg intake ratio | 3.9±1.5 | 3.7±0.9 | 0.32<sup>1</sup> |

Continuous variables: X±SD; categorical variables: %
Tests used: <sup>1</sup>Wilcoxon test; <sup>2</sup>Pearson chi-square test
eGFR: estimated glomerular filtration rate

The mean daily dose of personalized Mg supplementation was 205.59 mg with a range from 77.25 mg to 389.55 mg. Compliance with the pill regimen was very high for both the placebo and treatment arms (mean (standard deviation) based on pill counts were 96.1% (8.3) and 95.9% (10.2), respectively, and *P*=0.37 for difference between the arms). The mean Ca:Mg ratios (standard deviations) for the treatment and placebo arms after administering Mg and placebo supplementation were 2.27 (0.13) and 3.87 (1.46) respectively (*P* for difference, <0.001), based on the two 24-hour dietary recalls performed at baseline and remained stable at 2.13 (0.68) and 3.51 (1.32), respectively (*P* for difference, <0.001) based on the four 24-hour dietary recalls conducted over the 12-week period of the trial.

Shown in **Supplemental Table S1 and Table S2** are the effects of personalized Mg treatment on cytosine modifications 5-mC and 5-hmC, respectively, in the *TMPRSS2* for 32 CpG sites. We found, compared to the placebo arm, Mg treatment significantly increased 5-mC level at cg16371860 CpG site (*P*=0.011), even after adjusting for age and sex (*P*=0.010), and further adjusting for baseline 5-mC levels (*P*=0.019) (**Table 2**). However, the significance disappeared after false discovery rate (FDR) correction at 0.10 based on Benjamini–Hochberg approach. In stratified analyses by age (**Table 2**), we found that personalized Mg treatment significantly increased 5-mC methylation at cg16371860 (TSS1500, promoter) by 7.2% compared to placebo arm (decreased by 0.1%) in those aged < 65 years old. The difference remained statistically significant after adjusting for age, sex and baseline methylation as well as FDR correction (FDR-adjusted *P* =0.014). The effect of Mg treatment was not significant among those aged ≥ 65 years. On the other hand, we found that Mg treatment affected 5-hmC level at cg16371860 compared to the placebo arm after adjusting for age, sex, but the effect was not significant after further adjusting for baseline levels. In the stratified analysis by age, among those aged < 65 years, the effect of Mg treatment remained after adjusting for sex, age and baseline levels, but disappeared at FDR of 0.10.

**Table 2.** Changes in Cytosine Modifications (CpG sites) in *TMPRSS2* by Mg Treatment vs Placebo Stratified by Age

| CpG sites |  |  | Changes from baseline |  |  |  |  |  | P1 | P2 | P3 | FDR1 | FDR2 | FDR3 |
| --- | --- | --- | --- | --- | --- | --- | --- | --- | --- | --- | --- | --- | --- | --- |
|  |  |  | Placebo |  |  | Treatment |  |  |  |  |  |  |  |  |
|  |  |  | mean(SD) | median(25%, 75%) | % change in mean | mean(SD) | median(25%, 75%) | % change in mean |  |  |  |  |  |  |
| cg16371860 | 5-mC | Overall | -0.001(0.025) | -0.003(-0.014, 0.014) | 0.2 | 0.007(0.024) | 0.006(-0.006, 0.02) | 4.5 | 0.011 | 0.010 | 0.019 | 0.121 | 0.124 | 0.271 |
|  |  | Aged < 65 | -0.002(0.022) | -0.002(-0.012, 0.013) | -0.1 | 0.012(0.022) | 0.011(-0.001, 0.022) | 7.2 | 0.000 | 0.000 | 0.000 | 0.007 | 0.007 | 0.014 |
|  |  | Aged ≥ 65 | -0.001(0.031) | -0.003(-0.020, 0.015) | 0.7 | -0.004(0.025) | -0.004(-0.017, 0.008) | -1.6 | 0.650 | 0.735 | 0.633 | 0.951 | 0.930 | 0.981 |
|  | 5-hmC | Overall | 0.000(0.002) | 0.000(-0.001, 0.002) | 10.8 | -0.001(0.002) | -0.001(-0.001, 0.001) | -1.4 | 0.001 | 0.003 | 0.071 | 0.037 | 0.049 | 0.849 |
|  |  | Aged < 65 | 0.000(0.002) | 0.000(-0.001, 0.002) | 10.9 | -0.001(0.002) | -0.001(-0.001, 0.001) | -3.7 | 0.002 | 0.002 | 0.014 | 0.044 | 0.047 | 0.218 |
|  |  | Aged ≥ 65 | 0.000(0.002) | 0.001(-0.001, 0.002) | 10.5 | -0.000(0.002) | -0.000(-0.001, 0.001) | 3.8 | 0.325 | 0.488 | 0.423 | 0.743 | 0.958 | 0.883 |
| cg26337277 | 5-mC | Overall | 0.000(0.002) | 0.000(-0.001, 0.000) | NA | 0.000(0.001) | 0.000(0.000, 0.000) | NA | 0.759 | 0.827 | 0.325 | 0.899 | 0.913 | 0.743 |
|  |  | Aged < 65 | -0.000(0.002) | 0.000(-0.001, 0.000) | NA | -0.000(0.002) | 0.000(-0.000, 0.000) | NA | 0.678 | 0.676 | 0.991 | 0.868 | 0.865 | 0.991 |
|  |  | Aged ≥ 65 | 0.000(0.002) | 0.000(-0.001, 0.001) | NA | 0.000(0.001) | 0.000(0.000, 0.000) | NA | 0.275 | 0.310 | 0.143 | 0.874 | 0.919 | 0.943 |
|  | 5-hmC | Overall | 0.000(0.001) | 0.000(-0.001, 0.001) | 6.1 | -0.000(0.001) | -0.000(-0.001, 0.000) | -1.8 | 0.002 | 0.003 | 0.012 | 0.037 | 0.049 | 0.386 |
|  |  | Aged < 65 | 0.000(0.001) | 0.000(-0.001, 0.001) | 7.1 | -0.000(0.001) | -0.000(-0.001, 0.000) | -2.3 | 0.003 | 0.003 | 0.003 | 0.044 | 0.047 | 0.088 |
|  |  | Aged ≥ 65 | 0.000(0.001) | 0.000(-0.000, 0.001) | 3.9 | -0.000(0.001) | -0.000(-0.001, 0.000) | -0.6 | 0.302 | 0.358 | 0.979 | 0.743 | 0.969 | 0.844 |

Mg: magnesium;
*P*1: p value for crude GLM model; *P*2: p value for GLM model adjusting for age and sex; *P*3: p value for GLM model adjusting for age, sex and baseline methylation;
FDR: false discovery rate 0.10;
NA: % change in mean was not available due to extremely low levels of pre-treatment methylation.

Additionally, we found that Mg treatment reduced 5-hmC at cg26337277 (close proximity to TSS200 and 5’UTR, promoter) compared to placebo arm after adjusting for age, sex and baseline levels (*P*=0.012). However, the significant effect disappeared at FDR of 0.10 (FDR-adjusted *P*=0.386). In the stratified analysis by age, we found that Mg treatment significantly reduced 5-hmC level by 2.3% compared to increases by 7.1% in the placebo arm after adjusting for covariates in those aged < 65 years old (*P*=0.003). The effect remained significant at FDR of 0.10 (adjusted *P* value=0.088). No effect was found in those aged ≥ 65 years old.

## DISCUSSION and CONCLUSION

In this personalized precision-based randomized trial, we found that 12-week of Mg treatment significantly increased 5-mC DNA methylation at cg16371860 CpG site and decreased 5-hmC cytosine modification at cg26337277 CpG site in the *TMPRSS2* compared to placebo among participants aged less than 65 years old. To our best knowledge, no study has evaluated how to modify *TMPRSS2* cytosine modification, nor examined the effect of Mg treatment on the cytosine modification in the *TMPRSS2*.

Cg16371860 is a CpG site located at a CpG-rich island 200–1500 bp upstream of the transcriptional start site (TSS1500) while cg26337277 CpG site resides with close proximity to 0–200 bp upstream of the TSS (TSS200) and the 5’-untranslated region (5’UTR). These loci are within the promoter region of the gene, therefore potentially playing a role in transcription initiation and regulation of gene expression. Although 70-80% CpG sites of human genome are methylated to maintain a stable molecular phenotype, regions of CpG island promoters of actively transcribed genes are frequently lowly methylated [52]. Hypermethylation of the promoter regions could impede transcription activity and repress related gene expression [53]. Similarly, reduced 5-hmC level is also associated with decreased gene expression because conversion from 5-mC to 5-hmC is an active demethylation process [54]. Our observations that Mg treatment induced increases in 5-mC methylation at cg16371860 and decreases in 5-hmC methylation at cg26337277 CpG in the promoter indicate a hindered process of transcription initiation and, subsequently, lower levels of *TMPRSS2* expression.

Given that *TMPRSS2* plays an essential role in facilitating SARS-CoV-2 entry, higher expression of *TMPRSS2* may relate to higher viral loads in the host and subsequently, worse clinical outcomes. This has been supported by a recent finding that African Americans, individuals that were disproportionately affected by severe COVID-19 [55], had significantly higher average nasal epithelial gene expression of *TMPRSS2* compared with other races/ethnicities [56]. High viral loads can further induce cytokine storm which triggers a violent inflammatory immune response that contributes to acute respiratory distress syndrome (ARDS), multiple organ failure, and finally death in severe cases of SARS-CoV-2 infection [57,58]. The association between high viral loads and fatal clinical outcomes has been observed in both human influenza A (H5N1) [59] and SARS-CoV-2 [60].

In addition to viral entry, *TMPRSS2* expression levels may affect disease severity by regulating coagulation cascade and arterial thrombosis through the protease activated receptor (PAR)-signaling pathway [24–26]. This is further supported by a recent study reporting that polymorphisms near the *TMPRSS2* were associated with thrombocytes count [25]. Moreover, nafamostat [61], one *TMPRSS2* inhibitor has already been used in clinical practice as an effective anti-coagulant, and camostat mesylate [21], a closely related compound that are undergoing clinical trials to test their utility for COVID-19 treatment, further indicating the important role of *TMPRSS2* in regulating thrombosis and, in turn, disease severity. Given that nearly half of patients with COVID-19 pneumonia developed thrombotic complications [28] and deceased patients were distinctively characterized with widespread vascular thrombosis [27], *TMPRSS2* could be a key target for interventional strategies in reducing COVID-19 severity and mortality. Since TMPRSS2 plays a similar role during influenza infection [22], the findings from the current study suggest Mg status may also affect influenza’s infection and severity.

Strikingly, *TMPRSS2* was first identified as a key regulator in prostate cancer. Strongly upregulated *TMPRSS2* expression was observed in prostate cancer cell lines [62]. Consistent with racial disparities in nasal epithelial gene expression of *TMPRSS2*, African Americans also have higher incidence and mortality of prostate cancer than European Americans [63], further indicating that the high prostate cancer mortality among African Americans may be attributed to upregulated *TMPRSS2* expression. However, factors that may impact modified *TMPRSS2* expression remain unknown. Previous evidence found that African Americans had significantly lower levels of serum Mg or lower intake of Mg compared to European Americans [64] while low blood Mg levels and a high Ca/Mg ratio were significantly associated with high-grade prostate cancer [65]. Our findings from the current study in which Mg status was improved by modulating Ca: Mg intake ratio to around 2.3 suggest a protective effect from severe COVID-19 by Mg among individuals aged younger than 65 years through regulating DNA methylation/demethylation modifications and subsequently suppressing *TMPRSS2* expression.

Although no significant effect on *TMPRSS2* methylation was observed among those aged 65 years or older, it is likely Mg treatment may exert benefits through other mechanisms. First, our and other studies found Mg optimized body vitamin D status and substantially reduced the doses requirement for vitamin D [9,18,66]. Vitamin D has been proposed to generate beneficial effects in ARDS through activation of the vitamin D receptor (VDR) signaling pathway by reducing the cytokine storm, regulating the renin-angiotensin system, maintaining the integrity of the pulmonary epithelial barrier, and tapering down the increased coagulability [16]. Secondly, Mg deficiency is a prevailing, yet under-recognized driver for increased risks of cardiometabolic diseases including diabetes [67], hypertension, coronary heart disease, heart failure and thrombosis [68]. Previous studies found that low Mg plays an essential role in promoting endothelial cell dysfunction and generating a proinflammatory, pro-thrombotic and pro-atherogenic environment that could contribute to the pathogenesis of cardiovascular diseases and severe COVID-19 [69]. Thirdly, Mg deficiency caused by a gene mutation led to reduced cytotoxic activity of T cells and increased viral load, but Mg treatment reduced B cells positive for EBV, indicating Mg is critical in antivirus immunity [70]. Lastly, a study found that COVID-19 patients who carry the apolipoprotein E (*ApoE*) genotype were at a 2.3-fold increased risk of severe COVID-19 [71]. In addition to being a major risk factor for dementia [72], the *ApoE* genotype is strongly linked to lower human longevity [73], a genetic mark of early aging and is linked to lower plasma ApoE [74]. We reported from a randomized trial that in those age ≥65 years, Mg treatment improved cognitive function, particularly among elderly via demethylation in the *ApoE* gene which is expected to result in increased ApoE levels [47].

This study has several strengths, including the double-blinded randomized trial design. Furthermore, a precision-based design was used. Intakes of Mg and Ca from both diet and supplements were measured twice before and four times during the treatment and a personalized dosing strategy of Mg supplementation was administered to each participant. The Ca:Mg ratios remained stable over the 12-week study period. In addition, we had a high compliance with the study medication and the dropout rate was very low. The study has some weaknesses though. The primary concern is that *TMPRSS2* expression was not measured in the PPCCT, and thus the association between DNA methylation changes and level of *TMPRSS2* expression and phenotype patterns is not confirmed. However, the observed DNA methylation changes were internally consistent, with increased 5-mC and reduced 5-hmC indicating a reduced level of *TMPRSS2* expression.

In summary, among individuals aged younger than 65 years with the Ca:Mg intake ratios equal to or over 2.6, reducing Ca:Mg ratios to around 2.3 increased 5-mC modifications (i.e. cg16371860) and reduced 5-hmC modifications (i.e. cg26337277) in the *TMPRSS2* gene. The NIH has recently highlighted the crucial need for early intervention to reduce the likelihood of developing severe outcomes and reduce demand on healthcare system [1]. These findings, if confirmed, provide another mechanism for the role of Mg intervention for the prevention of COVID-19 and treatment of early and mild disease by modifying the phenotype of the *TMPRSS2* genotype in addition to affecting *ApoE* methylation in the elderly [47] and optimizing the levels of vitamin D [9]. In addition, these findings also indicate a possible mechanism of Mg involved in prostate cancer and influenza.

## Data Availability

The data have been deposited to the NIH database of Genotypes and Phenotypes (dbGaP).

## Author contribution

Lei Fan: Conceptualization, Formal analysis, Writing- Original draft preparation; Xiangzhu Zhu: Data curation, Methodology; Yinan Zheng: Investigation, Writing- Reviewing and Editing; Wei Zhang: Investigation, Writing- Reviewing and Editing; Douglas L Seidner: Supervision, Writing- Reviewing and Editing; Reid Ness: Supervision, Writing- Reviewing and Editing; Harvey J Murff: Supervision, Writing- Reviewing and Editing; Chang Yu: Methodology, Funding acquisition, Writing- Reviewing and Editing; Xiang Huang: Validation, Formal analysis; Martha J Shrubsole: Supervision, Writing- Reviewing and Editing; Lifang Hou: Supervision, Funding acquisition, Writing- Reviewing and Editing; Qi Dai: Conceptualization, Project administration, Funding acquisition, Writing- Reviewing and Editing.

## References

[1] Kim PS, Read SW, Fauci AS. Therapy for Early COVID-19: A Critical Need. JAMA 2020. https://doi.org/10.1001/jama.2020.22813.

[2] Wang Y, Zhang D, Du G, Du R, Zhao J, Jin Y, et al. Remdesivir in adults with severe COVID-19: a randomised, double-blind, placebo-controlled, multicentre trial. Lancet 2020;395:1569–78. https://doi.org/10.1016/S0140-6736(20)31022-9.

[3] Goldman JD, Lye DCB, Hui DS, Marks KM, Bruno R, Montejano R, et al. Remdesivir for 5 or 10 Days in Patients with Severe Covid-19. N Engl J Med 2020. https://doi.org/10.1056/NEJMoa2015301.

[4] Wilkinson E. RECOVERY trial: the UK covid-19 study resetting expectations for clinical trials. BMJ 2020;369:m1626. https://doi.org/10.1136/bmj.m1626.

[5] Beigel JH, Tomashek KM, Dodd LE, Mehta AK, Zingman BS, Kalil AC, et al. Remdesivir for the Treatment of Covid-19 - Final Report. N Engl J Med 2020. https://doi.org/10.1056/NEJMoa2007764.

[6] Rondy M, El Omeiri N, Thompson MG, Levêque A, Moren A, Sullivan SG. Effectiveness of influenza vaccines in preventing severe influenza illness among adults: A systematic review and meta-analysis of test-negative design case-control studies. J Infect 2017;75:381–94. https://doi.org/10.1016/j.jinf.2017.09.010.

[7] Korber B, Fischer WM, Gnanakaran S, Yoon H, Theiler J, Abfalterer W, et al. Tracking Changes in SARS-CoV-2 Spike: Evidence that D614G Increases Infectivity of the COVID-19 Virus. Cell 2020;182:812-827.e19. https://doi.org/10.1016/j.cell.2020.06.043.

[8] Li Q, Wu J, Nie J, Zhang L, Hao H, Liu S, et al. The Impact of Mutations in SARS-CoV-2 Spike on Viral Infectivity and Antigenicity. Cell 2020;182:1284-1294.e9. https://doi.org/10.1016/j.cell.2020.07.012.

[9] Dai Q, Zhu X, Manson JE, Song Y, Li X, Franke AA, et al. Magnesium status and supplementation influence vitamin D status and metabolism: results from a randomized trial. Am J Clin Nutr 2018;108:1249–58. https://doi.org/10.1093/ajcn/nqy274.

[10] Vázquez-Lorente H, Herrera-Quintana L, Molina-López J, Gamarra-Morales Y, López-González B, Miralles-Adell C, et al. Response of Vitamin D after Magnesium Intervention in a Postmenopausal Population from the Province of Granada, Spain. Nutrients 2020;12. https://doi.org/10.3390/nu12082283.

[11] Deng X, Song Y, Manson JE, Signorello LB, Zhang SM, Shrubsole MJ, et al. Magnesium, vitamin D status and mortality: results from US National Health and Nutrition Examination Survey (NHANES) 2001 to 2006 and NHANES III. BMC Med 2013;11:187. https://doi.org/10.1186/1741-7015-11-187.

[12] Tan CW, Ho LP, Kalimuddin S, Cherng BPZ, Teh YE, Thien SY, et al. Cohort study to evaluate the effect of vitamin D, magnesium, and vitamin B12 in combination on progression to severe outcomes in older patients with coronavirus (COVID-19). Nutrition 2020;79–80:111017. https://doi.org/10.1016/j.nut.2020.111017.

[13] Ali N. Role of vitamin D in preventing of COVID-19 infection, progression and severity. J Infect Public Health 2020. https://doi.org/10.1016/j.jiph.2020.06.021.

[14] Hastie C.E., Mackay D.F., Ho F., Celis-Morales C.A., Katikireddi S.V., Niedzwiedz C.L., Jani B.D., Welsh P., Mair F.S., Gray S.R., O’Donnell C.A., Gill J.M.R., Sattar N., Pell J.P. Vitamin D concentrations and COVID-19 infection in UK Biobank. Diabetes Metab. Syndr. Clin. Res. Rev. 2020;14:561–565. doi: 10.1016/j.dsx.2020.04.050.n.d.

[15] Rizzoli R. Vitamin D supplementation: upper limit for safety revisited? Aging Clin Exp Res 2020. https://doi.org/10.1007/s40520-020-01678-x.

[16] Entrenas Castillo M, Entrenas Costa LM, Vaquero Barrios JM, Alcalá Díaz JF, López Miranda J, Bouillon R, et al. Effect of calcifediol treatment and best available therapy versus best available therapy on intensive care unit admission and mortality among patients hospitalized for COVID-19: A pilot randomized clinical study. J Steroid Biochem Mol Biol 2020;203:105751. https://doi.org/10.1016/j.jsbmb.2020.105751.

[17] Reddy V, Sivakumar B. Magnesium-dependent vitamin-D-resistant rickets. Lancet 1974;1:963–5. https://doi.org/10.1016/s0140-6736(74)91265-3.

[18] Rude RK, Adams JS, Ryzen E, Endres DB, Niimi H, Horst RL, et al. Low serum concentrations of 1,25-dihydroxyvitamin D in human magnesium deficiency. J Clin Endocrinol Metab 1985;61:933–40. https://doi.org/10.1210/jcem-61-5-933.

[19] Fuss M, Bergmann P, Bergans A, Bagon J, Cogan E, Pepersack T, et al. Correction of low circulating levels of 1,25-dihydroxyvitamin D by 25-hydroxyvitamin D during reversal of hypomagnesaemia. Clin Endocrinol (Oxf) 1989;31:31–8. https://doi.org/10.1111/j.1365-2265.1989.tb00451.x.

[20] Ervin RB. Dietary Intake of Selected Minerals for the United States Population: 1999– 2000 2004:6.

[21] Hoffmann M, Kleine-Weber H, Schroeder S, Krüger N, Herrler T, Erichsen S, et al. SARS-CoV-2 Cell Entry Depends on ACE2 and TMPRSS2 and Is Blocked by a Clinically Proven Protease Inhibitor. Cell 2020;181:271-280.e8. https://doi.org/10.1016/j.cell.2020.02.052.

[22] Hatesuer B, Bertram S, Mehnert N, Bahgat MM, Nelson PS, Pöhlmann S, et al. Tmprss2 is essential for influenza H1N1 virus pathogenesis in mice. PLoS Pathog 2013;9:e1003774. https://doi.org/10.1371/journal.ppat.1003774.

[23] Asselta R, Paraboschi EM, Mantovani A, Duga S. ACE2 and TMPRSS2 variants and expression as candidates to sex and country differences in COVID-19 severity in Italy. Aging (Albany NY) 2020;12:10087–98. https://doi.org/10.18632/aging.103415.

[24] Isermann B. Homeostatic effects of coagulation protease-dependent signaling and protease activated receptors. J Thromb Haemost 2017;15:1273–84. https://doi.org/10.1111/jth.13721.

[25] Lopera Maya EA, van der Graaf A, Lanting P, van der Geest M, Fu J, Swertz M, et al. Lack of Association Between Genetic Variants at ACE2 and TMPRSS2 Genes Involved in SARS-CoV-2 Infection and Human Quantitative Phenotypes. Front Genet 2020;11:613. https://doi.org/10.3389/fgene.2020.00613.

[26] Sidhu TS, French SL, Hamilton JR. Differential signaling by protease-activated receptors: implications for therapeutic targeting. Int J Mol Sci 2014;15:6169–83. https://doi.org/10.3390/ijms15046169.

[27] Ackermann M, Verleden SE, Kuehnel M, Haverich A, Welte T, Laenger F, et al. Pulmonary Vascular Endothelialitis, Thrombosis, and Angiogenesis in Covid-19. N Engl J Med 2020;383:120–8. https://doi.org/10.1056/NEJMoa2015432.

[28] Klok FA, Kruip MJHA, van der Meer NJM, Arbous MS, Gommers D, Kant KM, et al. Confirmation of the high cumulative incidence of thrombotic complications in critically ill ICU patients with COVID-19: An updated analysis. Thromb Res 2020;191:148–50. https://doi.org/10.1016/j.thromres.2020.04.041.

[29] Dunn BK. Hypomethylation: one side of a larger picture. Ann N Y Acad Sci 2003;983:28–42. https://doi.org/10.1111/j.1749-6632.2003.tb05960.x.

[30] Robertson KD, Wolffe AP. DNA methylation in health and disease. Nat Rev Genet 2000;1:11–9. https://doi.org/10.1038/35049533.

[31] Jiangbo Wei, Alana Beadell, Zhou Zhang, Geeta Sharma, Raman Talwar, Patrick Arensdorf, Jason Karpus, Ajay Goel, Marc Bissonnette, Wei Zhang, Samuel Levy, Chuan He. A human tissue map of 5-hydroxymethylcytosines exhibits tissue specificity through gene and enhancer modulation. 2020 Nat Commun; In Press n.d.

[32] Jin S-G, Wu X, Li AX, Pfeifer GP. Genomic mapping of 5-hydroxymethylcytosine in the human brain. Nucleic Acids Res 2011;39:5015–24. https://doi.org/10.1093/nar/gkr120.

[33] Mariani CJ, Madzo J, Moen EL, Yesilkanal A, Godley LA. Alterations of 5-hydroxymethylcytosine in human cancers. Cancers (Basel) 2013;5:786–814. https://doi.org/10.3390/cancers5030786.

[34] Bishop KS, Ferguson LR. The interaction between epigenetics, nutrition and the development of cancer. Nutrients 2015;7:922–47. https://doi.org/10.3390/nu7020922.

[35] Ryu M-S, Langkamp-Henken B, Chang S-M, Shankar MN, Cousins RJ. Genomic analysis, cytokine expression, and microRNA profiling reveal biomarkers of human dietary zinc depletion and homeostasis. Proc Natl Acad Sci USA 2011;108:20970–5. https://doi.org/10.1073/pnas.1117207108.

[36] Mithieux G, Vega FV, Riou JP. The liver glucose-6-phosphatase of intact microsomes is inhibited and displays sigmoid kinetics in the presence of alpha-ketoglutarate-magnesium and oxaloacetate-magnesium chelates. J Biol Chem 1990;265:20364–8.

[37] Panov A, Scarpa A. Independent modulation of the activity of alpha-ketoglutarate dehydrogenase complex by Ca2+ and Mg2+. Biochemistry 1996;35:427–32. https://doi.org/10.1021/bi952101t.

[38] Lu X, Zhao BS, He C. TET family proteins: oxidation activity, interacting molecules, and functions in diseases. Chem Rev 2015;115:2225–39. https://doi.org/10.1021/cr500470n.

[39] Klungland A, Robertson AB. Oxidized C5-methyl cytosine bases in DNA: 5-Hydroxymethylcytosine; 5-formylcytosine; and 5-carboxycytosine. Free Radic Biol Med 2017;107:62–8. https://doi.org/10.1016/j.freeradbiomed.2016.11.038.

[40] Liu S, Liu Q. Personalized magnesium intervention to improve vitamin D metabolism: applying a systems approach for precision nutrition in large randomized trials of diverse populations. Am J Clin Nutr 2018;108:1159–61. https://doi.org/10.1093/ajcn/nqy294.

[41] Rosanoff A, Dai Q, Shapses SA. Essential Nutrient Interactions: Does Low or Suboptimal Magnesium Status Interact with Vitamin D and/or Calcium Status? Adv Nutr 2016;7:25–43. https://doi.org/10.3945/an.115.008631.

[42] Dai Q, Shu X-O, Deng X, Xiang Y-B, Li H, Yang G, et al. Modifying effect of calcium/magnesium intake ratio and mortality: a population-based cohort study. BMJ Open 2013;3. https://doi.org/10.1136/bmjopen-2012-002111.

[43] Zhao J, Giri A, Zhu X, Shrubsole MJ, Jiang Y, Guo X, et al. Calcium: magnesium intake ratio and colorectal carcinogenesis, results from the prostate, lung, colorectal, and ovarian cancer screening trial. Br J Cancer 2019;121:796–804. https://doi.org/10.1038/s41416-019-0579-2.

[44] Dai Q, Shrubsole MJ, Ness RM, Schlundt D, Cai Q, Smalley WE, et al. The relation of magnesium and calcium intakes and a genetic polymorphism in the magnesium transporter to colorectal neoplasia risk. Am J Clin Nutr 2007;86:743–51. https://doi.org/10.1093/ajcn/86.3.743.

[45] Nazor KL, Boland MJ, Bibikova M, Klotzle B, Yu M, Glenn-Pratola VL, et al. Application of a low cost array-based technique - TAB-Array - for quantifying and mapping both 5mC and 5hmC at single base resolution in human pluripotent stem cells. Genomics 2014;104:358–67. https://doi.org/10.1016/j.ygeno.2014.08.014.

[46] Zeng C, Zhang Z, Wang J, Chiu BC-H, Hou L, Zhang W. Application of the High-throughput TAB-Array for the Discovery of Novel 5-Hydroxymethylcytosine Biomarkers in Pancreatic Ductal Adenocarcinoma. Epigenomes 2019;3. https://doi.org/10.3390/epigenomes3030016.

[47] Zhu X, Borenstein AR, Zheng Y, Zhang W, Seidner DL, Ness R, et al. Ca:Mg Ratio, APOE Cytosine Modifications, and Cognitive Function: Results from a Randomized Trial. J Alzheimers Dis 2020;75:85–98. https://doi.org/10.3233/JAD-191223.

[48] Xu Z, Niu L, Li L, Taylor JA. ENmix: a novel background correction method for Illumina HumanMethylation450 BeadChip. Nucleic Acids Res 2016;44:e20. https://doi.org/10.1093/nar/gkv907.

[49] Tukey, John W. Exploratory data analysis. Vol. 2. 1977. n.d.

[50] Xu Z, Langie SAS, De Boever P, Taylor JA, Niu L. RELIC: a novel dye-bias correction method for Illumina Methylation BeadChip. BMC Genomics 2017;18:4. https://doi.org/10.1186/s12864-016-3426-3.

[51] Sanyaolu A, Okorie C, Marinkovic A, Patidar R, Younis K, Desai P, et al. Comorbidity and its Impact on Patients with COVID-19. SN Compr Clin Med 2020:1–8. https://doi.org/10.1007/s42399-020-00363-4.

[52] Greenberg MVC, Bourc’his D. The diverse roles of DNA methylation in mammalian development and disease. Nat Rev Mol Cell Biol 2019;20:590–607. https://doi.org/10.1038/s41580-019-0159-6.

[53] Abdolmaleky HM, Smith CL, Faraone SV, Shafa R, Stone W, Glatt SJ, et al. Methylomics in psychiatry: Modulation of gene-environment interactions may be through DNA methylation. Am J Med Genet B Neuropsychiatr Genet 2004;127B:51–9. https://doi.org/10.1002/ajmg.b.20142.

[54] Rodríguez-Aguilera JR, Ecsedi S, Goldsmith C, Cros M-P, Domínguez-López M, Guerrero-Celis N, et al. Genome-wide 5-hydroxymethylcytosine (5hmC) emerges at early stage of in vitro differentiation of a putative hepatocyte progenitor. Sci Rep 2020;10:7822. https://doi.org/10.1038/s41598-020-64700-2.

[55] Carethers JM. Insights into Disparities Observed with COVID-19. J Intern Med 2020. https://doi.org/10.1111/joim.13199.

[56] Bunyavanich S, Grant C, Vicencio A. Racial/Ethnic Variation in Nasal Gene Expression of Transmembrane Serine Protease 2 (TMPRSS2). JAMA 2020. https://doi.org/10.1001/jama.2020.17386.

[57] Nile SH, Nile A, Qiu J, Li L, Jia X, Kai G. COVID-19: Pathogenesis, cytokine storm and therapeutic potential of interferons. Cytokine Growth Factor Rev 2020;53:66–70. https://doi.org/10.1016/j.cytogfr.2020.05.002.

[58] Li X, Geng M, Peng Y, Meng L, Lu S. Molecular immune pathogenesis and diagnosis of COVID-19. J Pharm Anal 2020;10:102–8. https://doi.org/10.1016/j.jpha.2020.03.001.

[59] de Jong MD, Simmons CP, Thanh TT, Hien VM, Smith GJD, Chau TNB, et al. Fatal outcome of human influenza A (H5N1) is associated with high viral load and hypercytokinemia. Nat Med 2006;12:1203–7. https://doi.org/10.1038/nm1477.

[60] Magleby R, Westblade LF, Trzebucki A, Simon MS, Rajan M, Park J, et al. Impact of SARS-CoV-2 Viral Load on Risk of Intubation and Mortality Among Hospitalized Patients with Coronavirus Disease 2019. Clin Infect Dis 2020. https://doi.org/10.1093/cid/ciaa851.

[61] Choi J-Y, Kang Y-J, Jang HM, Jung H-Y, Cho J-H, Park S-H, et al. Nafamostat Mesilate as an Anticoagulant During Continuous Renal Replacement Therapy in Patients With High Bleeding Risk: A Randomized Clinical Trial. Medicine (Baltimore) 2015;94:e2392. https://doi.org/10.1097/MD.0000000000002392.

[62] Stopsack KH, Mucci LA, Antonarakis ES, Nelson PS, Kantoff PW. TMPRSS2 and COVID-19: Serendipity or Opportunity for Intervention? Cancer Discov 2020;10:779– 82. https://doi.org/10.1158/2159-8290.CD-20-0451.

[63] Mahal BA, Berman RA, Taplin M-E, Huang FW. Prostate Cancer-Specific Mortality Across Gleason Scores in Black vs Nonblack Men. JAMA 2018;320:2479–81. https://doi.org/10.1001/jama.2018.11716.

[64] Fowke JH, Koyama T, Dai Q, Zheng SL, Xu J, Howard LE, et al. Blood and dietary magnesium levels are not linked with lower prostate cancer risk in black or white men. Cancer Letters 2019;449:99–105. https://doi.org/10.1016/j.canlet.2019.02.023.

[65] Dai Q, Motley SS, Smith JA, Concepcion R, Barocas D, Byerly S, et al. Blood magnesium, and the interaction with calcium, on the risk of high-grade prostate cancer. PLoS ONE 2011;6:e18237. https://doi.org/10.1371/journal.pone.0018237.

[66] Cooper ID, Crofts CAP, DiNicolantonio JJ, Malhotra A, Elliott B, Kyriakidou Y, et al. Relationships between hyperinsulinaemia, magnesium, vitamin D, thrombosis and COVID-19: rationale for clinical management. Open Heart 2020;7. https://doi.org/10.1136/openhrt-2020-001356.

[67] Lima M de L, Pousada J, Barbosa C, Cruz T. [Magnesium deficiency and insulin resistance in patients with type 2 diabetes mellitus]. Arq Bras Endocrinol Metabol 2005;49:959–63. https://doi.org/10.1590/s0004-27302005000600016.

[68] DiNicolantonio JJ, O’Keefe JH, Wilson W. Subclinical magnesium deficiency: a principal driver of cardiovascular disease and a public health crisis. Open Heart 2018;5:e000668. https://doi.org/10.1136/openhrt-2017-000668.

[69] Maier JAM, Malpuech-Brugère C, Zimowska W, Rayssiguier Y, Mazur A. Low magnesium promotes endothelial cell dysfunction: implications for atherosclerosis, inflammation and thrombosis. Biochim Biophys Acta 2004;1689:13–21. https://doi.org/10.1016/j.bbadis.2004.01.002.

[70] Minton K. Immunodeficiency: magnesium regulates antiviral immunity. Nat Rev Immunol 2013;13:548–9. https://doi.org/10.1038/nri3501.

[71] Kuo C-L, Pilling LC, Atkins JL, Masoli JAH, Delgado J, Kuchel GA, et al. APOE e4 Genotype Predicts Severe COVID-19 in the UK Biobank Community Cohort. J Gerontol A Biol Sci Med Sci 2020;75:2231–2. https://doi.org/10.1093/gerona/glaa131.

[72] Michaelson DM. APOE ε4: the most prevalent yet understudied risk factor for Alzheimer’s disease. Alzheimers Dement 2014;10:861–8. https://doi.org/10.1016/j.jalz.2014.06.015.

[73] Sebastiani P, Gurinovich A, Nygaard M, Sasaki T, Sweigart B, Bae H, et al. APOE Alleles and Extreme Human Longevity. J Gerontol A Biol Sci Med Sci 2019;74:44–51. https://doi.org/10.1093/gerona/gly174.

[74] Rasmussen KL. Plasma levels of apolipoprotein E, APOE genotype and risk of dementia and ischemic heart disease: A review. Atherosclerosis 2016;255:145–55. https://doi.org/10.1016/j.atherosclerosis.2016.10.037.

