## Supplemental Table S1 and S2 for "Magnesium Treatment on Methylation Changes of Transmembrane Serine Protease 2 (TMPRSS2)"

| **Supplemental Table S1. Changes in 5-mC methylation (CpG sites) in TMPRSS2 by Mg treatment vs placebo** | | | | | | | | | |  |  |  |
| --- | --- | --- | --- | --- | --- | --- | --- | --- | --- | --- | --- | --- |
| **CpG sites** | **Changes from baseline** | | | | | | *P1* | *P2* | *P3* | FDR1 | FDR2 | FDR3 |
|  | **Placebo** | | | **Treatment** | | |  |  |  |  |  |  |
|  | mean(SD) | median(25%, 75%) | % change in mean | mean(SD) | median(25%, 75%) | % change in mean |  |  |  |  |  |  |
| cg02432628 | -0.003(0.016) | -0.002(-0.01, 0.006) | -0.3% | 0.002(0.016) | 0.001(-0.01, 0.014) | 0.2% | 0.0200 | 0.0200 | 0.0490 | 0.121 | 0.124 | 0.271 |
| cg16371860 | -0.001(0.025) | -0.003(-0.014, 0.014) | 0.2% | 0.007(0.024) | 0.006(-0.006, 0.02) | 4.5% | 0.0110 | 0.0100 | 0.0190 | 0.121 | 0.124 | 0.271 |
| cg07808966 | 0.002(0.029) | 0.002(-0.017, 0.018) | 0.3% | -0.002(0.03) | -0.002(-0.022, 0.02) | -0.1% | 0.3590 | 0.3360 | 0.3050 | 0.716 | 0.671 | 0.743 |
| cg26337277 | 0.000(0.002) | 0.000(-0.001, 0.000) | NA | 0.000(0.001) | 0.000(0.000, 0.000) | NA | 0.7590 | 0.8270 | 0.3250 | 0.899 | 0.913 | 0.743 |
| cg16084872 | -0.005(0.026) | -0.003(-0.019, 0.009) | -0.3% | 0.004(0.027) | 0.005(-0.013, 0.021) | 7.1% | 0.0230 | 0.0270 | 0.0510 | 0.121 | 0.124 | 0.271 |
| cg07919906 | -0.002(0.033) | -0.002(-0.019, 0.019) | -0.2% | 0.000(0.026) | -0.002(-0.02, 0.014) | 0.0% | 0.5960 | 0.5840 | 0.5110 | 0.829 | 0.779 | 0.849 |
| cg14982276 | 0.000(0.012) | 0.001(-0.006, 0.008) | 36.5% | -0.001(0.012) | -0.002(-0.008, 0.008) | 23.0% | 0.6590 | 0.7560 | 0.6900 | 0.879 | 0.890 | 0.849 |
| cg26309194 | -0.003(0.021) | -0.004(-0.014, 0.008) | -0.2% | -0.001(0.017) | 0.001(-0.009, 0.008) | 4.3% | 0.4340 | 0.3990 | 0.6900 | 0.77 | 0.709 | 0.849 |
| cg05218995 | -0.008(0.072) | -0.005(-0.043, 0.037) | -0.5% | 0.011(0.059) | 0.007(-0.027, 0.038) | 2.7% | 0.0460 | 0.0590 | 0.0660 | 0.183 | 0.234 | 0.301 |
| cg02702369 | -0.006(0.028) | -0.004(-0.022, 0.014) | -0.7% | -0.004(0.026) | -0.005(-0.022, 0.01) | -0.5% | 0.6870 | 0.5410 | 0.4780 | 0.88 | 0.768 | 0.849 |
| cg05563672 | -0.001(0.003) | 0.000(-0.001, 0.000) | NA | 0.000(0.002) | 0.000(0.000, 0.000) | NA | 0.2630 | 0.2360 | 0.9850 | 0.562 | 0.539 | 0.992 |
| cg15014277 | 0.000(0.006) | 0.000(-0.004, 0.003) | NA | 0.000(0.006) | 0.000(-0.003, 0.004) | NA | 0.3810 | 0.3590 | 0.4760 | 0.716 | 0.675 | 0.849 |
| cg02610779 | -0.006(0.037) | -0.008(-0.032, 0.018) | -0.8% | 0.007(0.039) | 0.002(-0.013, 0.025) | 1.5% | 0.0140 | 0.0150 | 0.0310 | 0.121 | 0.124 | 0.271 |
| cg19315202 | -0.002(0.007) | -0.002(-0.007, 0.003) | NA | 0.001(0.006) | 0.000(-0.003, 0.005) | NA | 0.0050 | 0.0040 | 0.0480 | 0.121 | 0.119 | 0.271 |
| cg02642997 | 0.002(0.022) | 0.000(-0.014, 0.014) | 12.7% | -0.002(0.027) | 0.001(-0.016, 0.014) | 19.7% | 0.2530 | 0.2260 | 0.4660 | 0.562 | 0.539 | 0.849 |
| cg00255189 | 0.002(0.023) | 0.003(-0.011, 0.014) | 0.3% | 0.003(0.024) | 0.005(-0.016, 0.021) | 0.4% | 0.8230 | 0.7600 | 0.8430 | 0.94 | 0.89 | 0.93 |
| cg00739644 | 0.002(0.055) | 0.000(-0.031, 0.032) | NA | -0.003(0.054) | -0.003(-0.03, 0.035) | NA | 0.5400 | 0.5520 | 0.7420 | 0.785 | 0.768 | 0.874 |
| cg19020860 | 0.002(0.06) | 0.001(-0.031, 0.028) | 0.7% | 0.002(0.052) | -0.002(-0.027, 0.034) | 0.6% | 0.9310 | 0.9280 | 0.3200 | 0.96 | 0.941 | 0.743 |
| cg22025068 | 0.001(0.032) | 0.000(-0.015, 0.025) | 2.1% | 0.008(0.031) | 0.007(-0.012, 0.024) | 5.0% | 0.1410 | 0.1510 | 0.2530 | 0.41 | 0.441 | 0.737 |
| cg18156003 | 0.002(0.03) | 0.004(-0.02, 0.018) | 0.3% | -0.004(0.03) | 0.000(-0.021, 0.014) | -0.4% | 0.1720 | 0.1820 | 0.9920 | 0.459 | 0.485 | 0.992 |
| cg00689211 | 0.000(0.014) | -0.001(-0.006, 0.006) | NA | 0.000(0.014) | 0.000(-0.007, 0.007) | NA | 0.9600 | 0.9410 | 0.6310 | 0.96 | 0.941 | 0.849 |
| cg12203615 | 0.000(0.043) | -0.001(-0.024, 0.017) | 9.1% | -0.002(0.039) | -0.002(-0.02, 0.018) | 9.9% | 0.7380 | 0.7790 | 0.5440 | 0.899 | 0.89 | 0.849 |
| cg12384236 | 0.000(0.011) | 0.000(-0.009, 0.008) | NA | 0.001(0.01) | 0.000(-0.006, 0.008) | NA | 0.4630 | 0.5370 | 0.2470 | 0.77 | 0.768 | 0.737 |
| cg13489049 | -0.001(0.027) | 0.001(-0.016, 0.017) | 0.1% | 0.008(0.028) | 0.006(-0.01, 0.029) | 2.5% | 0.0220 | 0.0270 | 0.0160 | 0.121 | 0.124 | 0.271 |
| cg13698049 | -0.003(0.03) | -0.002(-0.025, 0.018) | -0.3% | -0.004(0.03) | -0.005(-0.02, 0.012) | -0.3% | 0.9060 | 0.8630 | 0.9710 | 0.96 | 0.921 | 0.992 |
| cg16385367 | 0.004(0.039) | 0.008(-0.016, 0.029) | 0.7% | 0.001(0.035) | -0.002(-0.018, 0.019) | 0.2% | 0.4910 | 0.4700 | 0.4450 | 0.77 | 0.768 | 0.849 |
| cg01036509 | -0.002(0.011) | -0.002(-0.008, 0.007) | NA | -0.001(0.009) | -0.001(-0.008, 0.005) | NA | 0.9050 | 0.7510 | 0.7640 | 0.96 | 0.89 | 0.874 |
| cg24901042 | -0.005(0.027) | -0.002(-0.015, 0.01) | 0.5% | 0.001(0.024) | -0.001(-0.011, 0.015) | 11.6% | 0.0780 | 0.0960 | 0.2140 | 0.259 | 0.307 | 0.737 |
| cg01157146 | 0.003(0.059) | -0.001(-0.034, 0.04) | 1.4% | 0.008(0.057) | 0.014(-0.029, 0.037) | 2.3% | 0.5060 | 0.5160 | 0.6870 | 0.77 | 0.768 | 0.849 |
| cg02978411 | 0.006(0.024) | 0.004(-0.01, 0.02) | 0.8% | 0.001(0.021) | 0.000(-0.015, 0.012) | 0.1% | 0.0810 | 0.0780 | 0.6880 | 0.259 | 0.277 | 0.849 |
| cg02613803 | -0.001(0.018) | -0.001(-0.01, 0.009) | NA | 0.005(0.018) | 0.003(-0.007, 0.017) | NA | 0.0270 | 0.0230 | 0.0800 | 0.126 | 0.124 | 0.321 |
| cg19974120 | -0.001(0.022) | -0.001(-0.011, 0.012) | 0.0% | -0.004(0.019) | -0.005(-0.016, 0.005) | -0.4% | 0.2320 | 0.2590 | 0.5950 | 0.562 | 0.552 | 0.849 |
| Mg: magnesium; *P1*: p value for crude GLM model; *P2*: p value for GLM model adjusting for age and sex; *P3*: p value for GLM model adjusting for age, sex and baseline methylation; FDR: false discovery rate at 0.10;  NA: % change in mean was not available due to extremely low levels of pre-treatment methylation. | | | | | | | | | |  |  |  |

| **Supplemental Table S2. Changes in 5-hmC methylation (CpG sites) in TMPRSS2 by Mg treatment vs placebo** | | | | | | | | | |  |  |  |
| --- | --- | --- | --- | --- | --- | --- | --- | --- | --- | --- | --- | --- |
| **CpG sites** | **Changes from baseline** | | | | | |  |  |  |  |  |  |
|  | **Placebo** | | | **Treatment** | | |  |  |  |  |  |  |
|  | mean (SD) | median(25%, 75%) | % change in mean | mean (SD) | Median (25%, 75%) | % change in mean | *P1* | *P2* | *P3* | FDR1 | FDR2 | FDR3 |
| cg02432628 | -0.000(0.006) | -0.000(-0.004, 0.003) | 10.1% | 0.000(0.006) | -0.000(-0.003, 0.003) | 11.2% | 0.8802 | 0.8394 | 0.6413 | 0.9023 | 0.8960 | 0.9662 |
| cg16371860 | 0.000(0.002) | 0.000(-0.001, 0.002) | 10.8% | -0.001(0.002) | -0.001(-0.001, 0.001) | -1.4% | 0.0015 | 0.0031 | 0.0709 | 0.0367 | 0.0493 | 0.8489 |
| cg07808966 | -0.000(0.008) | 0.000(-0.003, 0.003) | 22.2% | -0.001(0.009) | -0.000(-0.004, 0.003) | 32.2% | 0.5389 | 0.6401 | 0.8792 | 0.8951 | 0.8921 | 0.9662 |
| cg26337277 | 0.000(0.001) | 0.000(-0.001, 0.001) | 6.1% | -0.000(0.001) | -0.000(-0.001, 0.000) | -1.8% | 0.0023 | 0.0026 | 0.0121 | 0.0367 | 0.0493 | 0.3859 |
| cg16084872 | 0.002(0.008) | 0.001(-0.003, 0.008) | 14.5% | 0.001(0.007) | 0.000(-0.004, 0.005) | 9.1% | 0.4422 | 0.4503 | 0.6966 | 0.8951 | 0.8921 | 0.9662 |
| cg07919906 | 0.002(0.016) | 0.002(-0.005, 0.011) | 23.9% | 0.001(0.016) | 0.000(-0.008, 0.008) | 19.8% | 0.5657 | 0.5371 | 0.2615 | 0.8951 | 0.8921 | 0.8489 |
| cg14982276 | -0.000(0.003) | -0.000(-0.002, 0.002) | 0.3% | -0.000(0.003) | 0.000(-0.002, 0.002) | 1.7% | 0.6220 | 0.6608 | 0.2544 | 0.8951 | 0.8921 | 0.8489 |
| cg26309194 | 0.000(0.002) | -0.000(-0.001, 0.001) | 2.5% | 0.000(0.002) | 0.000(-0.001, 0.001) | 3.9% | 0.5313 | 0.5450 | 0.3979 | 0.8951 | 0.8921 | 0.8489 |
| cg05218995 | -0.000(0.004) | -0.000(-0.003, 0.002) | 4.2% | 0.000(0.004) | 0.000(-0.002, 0.003) | 9.0% | 0.2451 | 0.2861 | 0.3734 | 0.8951 | 0.8921 | 0.8489 |
| cg02702369 | 0.001(0.015) | 0.001(-0.007, 0.009) | 22.5% | 0.001(0.014) | 0.002(-0.006, 0.009) | 21.0% | 0.8861 | 0.8466 | 0.5676 | 0.9023 | 0.8960 | 0.9662 |
| cg05563672 | 0.000(0.002) | -0.000(-0.001, 0.001) | 4.3% | 0.000(0.002) | 0.000(-0.001, 0.001) | 3.3% | 0.9023 | 0.8960 | 0.9371 | 0.9023 | 0.8960 | 0.9662 |
| cg15014277 | 0.000(0.003) | 0.000(-0.002, 0.002) | 3.8% | 0.000(0.004) | 0.001(-0.002, 0.003) | 5.4% | 0.6185 | 0.4387 | 0.3545 | 0.8951 | 0.8921 | 0.8489 |
| cg02610779 | 0.000(0.009) | 0.000(-0.005, 0.004) | 20.0% | -0.000(0.008) | -0.000(-0.004, 0.004) | 12.3% | 0.6392 | 0.6213 | 0.8472 | 0.8951 | 0.8921 | 0.9662 |
| cg19315202 | 0.000(0.003) | 0.000(-0.002, 0.002) | 4.9% | -0.000(0.004) | -0.000(-0.002, 0.002) | 1.1% | 0.2184 | 0.1947 | 0.3139 | 0.8951 | 0.8921 | 0.8489 |
| cg02642997 | 0.000(0.003) | -0.000(-0.001, 0.002) | 6.0% | -0.001(0.003) | -0.000(-0.003, 0.001) | -1.2% | 0.1339 | 0.1220 | 0.1459 | 0.8951 | 0.8921 | 0.8489 |
| cg00255189 | -0.000(0.012) | 0.000(-0.008, 0.006) | 2.1% | -0.001(0.012) | -0.001(-0.009, 0.008) | 1.4% | 0.7832 | 0.8458 | 0.3611 | 0.8951 | 0.8960 | 0.8489 |
| cg00739644 | 0.001(0.028) | 0.002(-0.018, 0.020) | 8.7% | -0.002(0.031) | 0.002(-0.020, 0.017) | 9.4% | 0.5249 | 0.5021 | 0.1048 | 0.8951 | 0.8921 | 0.8489 |
| cg19020860 | -0.003(0.046) | -0.002(-0.021, 0.017) | 21.1% | -0.001(0.042) | -0.001(-0.020, 0.016) | 25.9% | 0.6548 | 0.5857 | 0.9662 | 0.8951 | 0.8921 | 0.9662 |
| cg22025068 | -0.001(0.010) | 0.000(-0.006, 0.004) | 13.3% | -0.000(0.009) | 0.000(-0.004, 0.004) | 9.1% | 0.8823 | 0.8705 | 0.4879 | 0.9023 | 0.8960 | 0.9662 |
| cg18156003 | -0.001(0.019) | -0.001(-0.011, 0.010) | 10.1% | 0.002(0.020) | -0.000(-0.011, 0.011) | 19.4% | 0.3274 | 0.3148 | 0.7308 | 0.8951 | 0.8921 | 0.9662 |
| cg00689211 | 0.000(0.007) | 0.001(-0.003, 0.004) | 12.2% | -0.001(0.010) | -0.002(-0.007, 0.007) | 13.0% | 0.4151 | 0.4061 | 0.3195 | 0.8951 | 0.8921 | 0.8489 |
| cg12203615 | -0.000(0.004) | -0.000(-0.002, 0.002) | 2.7% | -0.000(0.004) | -0.001(-0.002, 0.002) | 1.4% | 0.7322 | 0.6969 | 0.8527 | 0.8951 | 0.8921 | 0.9662 |
| cg12384236 | 0.001(0.007) | -0.000(-0.003, 0.005) | 16.3% | 0.000(0.007) | 0.000(-0.004, 0.004) | 10.7% | 0.4398 | 0.5377 | 0.2724 | 0.8951 | 0.8921 | 0.8489 |
| cg13489049 | 0.001(0.012) | 0.001(-0.005, 0.008) | 8.4% | 0.000(0.013) | 0.001(-0.007, 0.007) | 10.9% | 0.7152 | 0.7311 | 0.7983 | 0.8951 | 0.8960 | 0.9662 |
| cg13698049 | 0.003(0.013) | 0.003(-0.005, 0.010) | 20.7% | 0.000(0.014) | 0.000(-0.008, 0.009) | 17.0% | 0.1828 | 0.1979 | 0.2831 | 0.8951 | 0.8921 | 0.8489 |
| cg16385367 | 0.000(0.008) | -0.000(-0.004, 0.003) | 15.7% | -0.001(0.008) | 0.000(-0.005, 0.004) | 7.9% | 0.3935 | 0.4227 | 0.6713 | 0.8951 | 0.8921 | 0.9662 |
| cg01036509 | 0.000(0.005) | 0.001(-0.003, 0.003) | 4.9% | 0.000(0.005) | 0.001(-0.004, 0.003) | 3.9% | 0.7575 | 0.6826 | 0.8170 | 0.8951 | 0.8921 | 0.9662 |
| cg24901042 | 0.000(0.008) | -0.000(-0.004, 0.005) | 8.3% | 0.001(0.008) | 0.001(-0.005, 0.005) | 12.6% | 0.6493 | 0.6839 | 0.3329 | 0.8951 | 0.8921 | 0.8489 |
| cg01157146 | -0.000(0.051) | 0.003(-0.034, 0.028) | 17.0% | -0.004(0.049) | -0.002(-0.032, 0.030) | 6.6% | 0.6144 | 0.6112 | 0.5253 | 0.8951 | 0.8921 | 0.9662 |
| cg02978411 | -0.001(0.007) | -0.001(-0.006, 0.004) | 2.6% | -0.002(0.008) | -0.001(-0.006, 0.005) | 1.6% | 0.6800 | 0.7835 | 0.9206 | 0.8951 | 0.8960 | 0.9662 |
| cg02613803 | 0.001(0.009) | 0.001(-0.005, 0.006) | 13.3% | -0.001(0.010) | -0.000(-0.006, 0.006) | 9.7% | 0.3514 | 0.2577 | 0.6201 | 0.8951 | 0.8921 | 0.9662 |
| cg19974120 | 0.000(0.017) | -0.001(-0.007, 0.006) | 29.4% | 0.002(0.015) | 0.002(-0.004, 0.010) | 37.4% | 0.4007 | 0.4238 | 0.6131 | 0.8951 | 0.8921 | 0.9662 |
| Mg: magnesium; *P1*: p value for crude GLM model; *P2*: p value for GLM model adjusting for age and sex; *P3*: p value for GLM model adjusting for age, sex and baseline methylation; FDR: false discovery rate at 0.10;  NA: % change in mean was not available due to extremely low levels of pre-treatment methylation. | | | | | | | | | |  |  |  |
